# Contribution of rare variants to heritability of a disease is much greater than conventionally estimated: modification of allele distribution model

**DOI:** 10.1101/2023.04.25.23289037

**Authors:** Yoshiro Nagao

## Abstract

Missing heritability is a current problem in human genetics. I previously reported a method to estimate heritability of a polymorphism (h_p_^2^) for a common disease without calculating the genetic variance under dominant and the recessive models. Here, I extended the method to the co-dominant model and carry out trial calculations of h_p_^2^. I also calculated h_p_^2^ applying the allele distribution model originally reported by Pawitan et al. for a comparison. Unexpectedly, h_p_^2^ calculated for rare variants with high odds ratios was much higher. I noticed that conventional methods use the allele frequency (AF) of a variant in the general population. However, this implicitly assumes that the unaffected are included among the phenotypes: an assumption that is inconsistent with case-control studies in which unaffected individuals belong to the control group. Therefore, I modified the allele distribution model by using the AF in the patient population. Consequently, the h_p_^2^ of rare variants was quite high. Recalculating h_p_^2^ of several rare variants reported in the literature with the modified allele distribution model, yielded results were 3.2 - 53.7 times higher than the original model. These results suggest that the contribution of rare variants to heritability of a disease has been considerably underestimated.

---

Genome-wide association studies (GWAS) for diseases are based on case-control studies^1^. GWAS have identified thousands of genetic polymorphisms associated with common diseases, however, every effort to account for more than a fraction of the heritability of the disease by the discovered variants has failed. This is called the missing heritability problem^2,3^.

Heritability (h^2^) is a concept that summarizes how much of the variation in a trait is due to variation in genetic factors^4^. The phenotypic variance in the trait (V_P_) is the sum of genetic variance (V_G_) and environmental variance (V_E_) as follows:

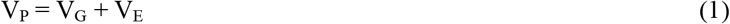

h^2^ is defined as:

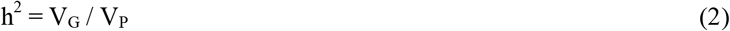

Considering a specific disease as a trait here, V_P_ and V_G_ represent the variations of phenotypes and genotypes of the disease in the population, respectively.

In conventional studies estimating the contribution of variants to h^2^ a co-dominant model is commonly assumed, and various estimation methods have been proposed^5,6^. Among them, the method of Pawitan et al. is simple. According to this formulation, when n variants are associated with a disease the variance of the risk distribution, V(g)_k_, for the k-th variant is represented by its allele frequency (AF) in the general population, p_k_, and odds ratio, OR_k_, by the following equation:

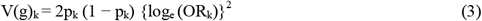

V(g)_k_ is considered as the genetic variance of the k-th variant, and there should be the following relation between V(g)_k_ and V_G_.

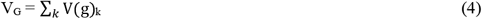

h^2^ is evaluated by the following equation:

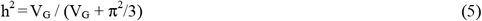

Oliynyk called the method using the equations (3), (4) and (5) the “allele distribution model” ^7^.

In a previous report, I presented a method to estimate h^2^ of a polymorphism (h_p_^2^) by probabilistically predicting how the risk genotype of a variant is inherited by the relatives of a patient under dominant and the recessive models^8^. In the present study, I extend the approach to the co-dominant model and do trial calculations of h_p_^2^ for variants with various ORs to assess the results compared to the standard method.

## Results

### Definitions and premises

- p is the AF of the risk allele of a variant in the general population
- q is the AF of the non-risk allele of a variant in the general population.
- u is the AF of the risk allele of a variant in the patient population.
- v is the AF of the non-risk allele of a variant in the patient population
- **P** is the Prevalence of a disease.
- **Q** is the recurrence risk of the disease among the first-degree relatives of a patient

### Derivation of the allele frequency of a variant in the patient population

If the frequencies of the risk allele and the non-risk allele in the unaffected group are x and y, respectively, the following equation holds^8^:

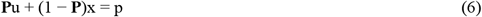

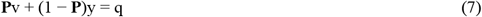

When OR is represented as k, the following equation holds:

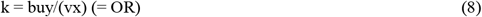

Simultaneous equations (8), (9) and (10) cannot be solved analytically, but they can be solved numerically using the following approximation:

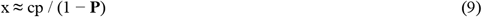

In this relation, c is a constant with a value between 1 and 0. The approximate solution for u then becomes

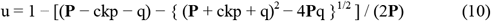

The value of c is estimated by checking the OR’. Allowing for an error of +/- 1 %, the value of OR’/OR should be between 0.99 and 1.01.

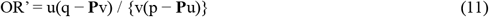

### Derivation of the equation for the “mutation model”

Assuming that a polymorphism (variant) is the only genetic factor for a disease, **Q** is represented, for a variant that acts in a co-dominant manner, by Equation (12):

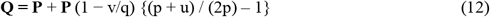

Once **Q** is estimated, h_p_^2^ is calculated by the Falconer liability threshold model^8^. [Details of the calculation are given in Falconer^9^.] Equation (12) assumes that the non-risk genotype has no genetic effect; in other words, the variant is regarded as a mutation. Indeed, substituting **P** = p and u = 1 into Equation (12) yields a **Q** of about 0.5, which is the incidence in first-degree relatives when the patient is homozygote for a Mendelian disease with autosomal co-dominant inheritance. Therefore, Equation (12) describes what can be called the “mutation model”.

### Application of the equation of the allele distribution model to calculate h_p_^2^

Equation (5) is a formula for to estimate the heritability of all variants taken together; however, it can also be used to estimate the contribution of individual variants to h^2^. The following equations are derived to estimate h_p_^2^ for a variant that has a MAF of p and an odds ratio of OR:

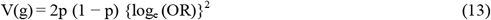

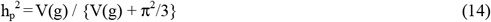

### Comparison of the h_p_^2^ calculated with the allele distribution model and with the mutation model

The calculated values of the h_p_^2^ by the two methods can now be compared. In Fig. 1, h_p_^2^ calculated by the allele distribution model (h_1_^2^) or by the mutation model (h_2_^2^), are shown for various values of OR (Figs.1a-1d). In the calculation of h_2_^2^, **P** was set to 0.01 (as in the case of schizophrenia, e.g., Schultz et al.^10^). h_1_^2^ generally has a greater value than h_2_^2^, however, when OR is high and AF is low, h_2_^2^ is greater than h_1_^2^. As shown in Fig. 1b, h_2_^2^ is always lower than h_1_^2^ when OR is 4, but when OR is 10, as shown in Fig. 1c, h_2_^2^ is higher than h_1_^2^ at low AF.

**FIGURE 1.**
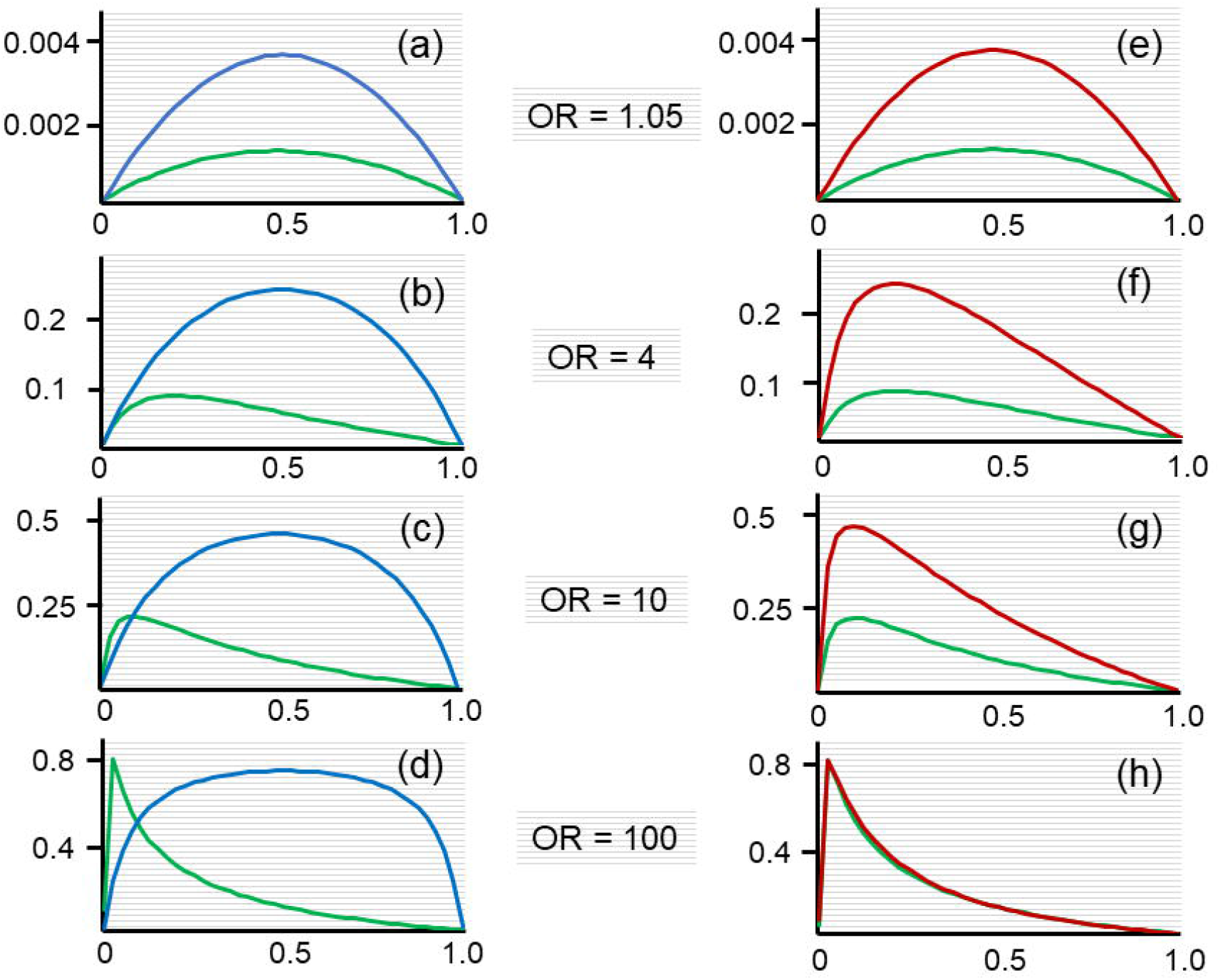
h_p_^2^ of variants calculated by three methods for various odds ratios. (a) to (d) h_p_^2^ of variants are shown for odds ratio (OR) of 1.05, 4, 10 and 100 calculated with the allele distribution model (blue line) and with the mutation model (green line). Vertical axis, value of h_p_^2^; horizontal axis, allele frequency in the general population. (e) to (h) h_p_^2^ of variants are shown for OR of 1.05, 4, 10 and 100 calculated with the modified allele distribution model (red line) and with the mutation model (green line). Vertical axis, value of h_p_^2^; horizontal axis, allele frequency in the general population.

Why are the calculated values of h_p_^2^ by the two methods so different? Looking at Equation (3), we can see that V(g)_k_ is calculated using the AF of a variant in the general population, p_k_. Therefore, V_G_ defined by Equation (4) is also the genetic variance of a disease in the general population. If Equation (2) holds, V_P_ should also be the phenotypic variance of a disease in the general population. But of course, the general population includes the large number of individuals who do not have the disease, and the conventional method has calculated V(g)_k_ of k-th variant assuming that the phenotypes of a disease include those of all the unaffected individuals. This assumption is, however, inconsistent with the established rule that in a case-control study, unaffected individuals belong to the control (unaffected) group. I therefore propose here that V_P_ in Equation (4) should be the phenotypic variance of a disease in the patient population. Therefore, V_G_ should be the genotypic variance in the patient population and the V(g)_k_ of the k-th variant should be calculated using the AF in the patient cohort.

### Derivation of the modified equation for the allele distribution model

Pawitan et al. assume that the proportion of SNPs associated with a certain disease is normally distributed when plotted against the log scale value of OR for each SNP on the x-axis (Fig. 2)^5^. In their model the mean value of polygenic risk score (β_mean_) in the general population is represented by the following equation^7^:

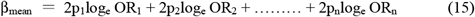

**FIGURE 2.**
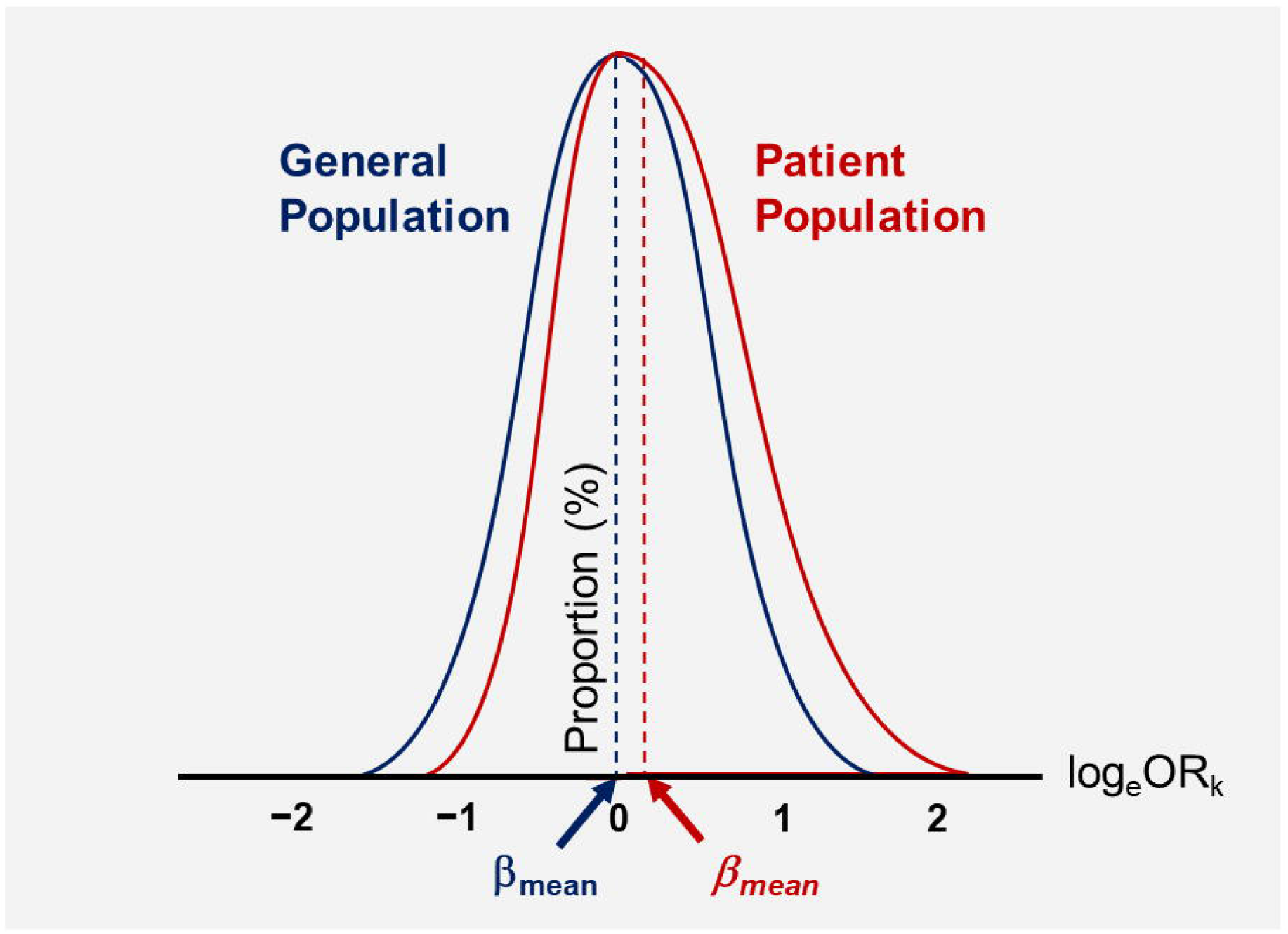
Distribution of the proportion of variants that are associated with a disease. The proportions of variants are normally distributed in the general population (blue line), whereas the distribution of variant proportions in the patient population becomes asymmetric (red line). β_mean_ measures the deviation of the mean value of log_e_ OR from y-axis in the general population and *β*_mean_ measures the deviation of the mean value of log_e_ OR from y-axis in the patient population. Horizontal axis: log scaled OR of variant Vertical axis: proportion of variant

In this equation, p_k_ and OR_k_ are minor allele frequency (MAF) and OR of the k-th SNP, respectively.

What is the expected for β_mean_ in the general population? Under the assumption that log_e_ OR for each SNP is normally distributed, it should be 0.

In a similar way, the mean value of polygenic risk score for patients of a disease (*β*_mean_) is represented by the following equation:

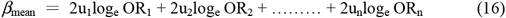

In this equation u_k_ and OR_k_ are MAF in the patient population and OR of the k-th SNP, respectively. In formula (16), for SNPs with positive log_e_ OR_k_, u_k_ is generally larger than p_k_, and u_k_ is generally smaller than p_k_ for SNPs with negative log_e_ OR_k_. Consequently, β_mean_ becomes greater than 0. Schematic images of β_mean_ and *β*_mean_ are represented in Fig. 2. Comparing Equations (15) and (16), V(g) of a variant in a patient population is represented as

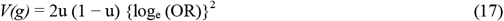

Therefore, h_p_^2^ is calculated as

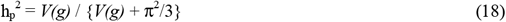

The method using Equations (17) and (18) can be termed the “modified allele distribution model”. In Equation (17), u is a function of p because it can be calculated using p, **P** and OR. In Fig. 1, h_p_^2^ calculated with the modified allele distribution model (h_3_^2^) is shown for various ORs in comparison with the h_p_^2^ calculated with the mutation model (h_2_^2^) (Figs. 1e-1h). h_3_^2^ is always higher than h_2_^2^, but the graphs are similar in shape. The curves for h_1_^2^ and h_3_^2^ look similar for low values of OR, but diverge as OR increases (Figs. 1a-1h).

What is the expected for β_mean_ in the general population? Under the assumption that log_e_ OR for each SNP is normally distributed, it should be 0.

### Comparison of h_p_^2^ by three methods for variants from the literature

Table 1 shows a comparison of h_p_^2^ assessed by the three methods discussed here for a number of single nucleotide polymorphisms (SNPs) and copy number variations (CNVs) cited from the literatures^11-24^. The values of h_3_^2^ for variants with common AFs and intermediate AFs (p ≥ 0.05) were not so different from h_1_^2^, while h_2_^2^ was obviously lower. For so-called rare variants with low AFs (p < 0.01), h_3_^2^ was the highest, and the difference from h_1_^2^ was prominent, with a ratio ranging from 3.2 up to 53.7-fold. All variants whose risk alleles were not found in the control group were scored as rare variants, and h_p_^2^ was calculated as p = **P**u using the mutation model.

**Table 1.**
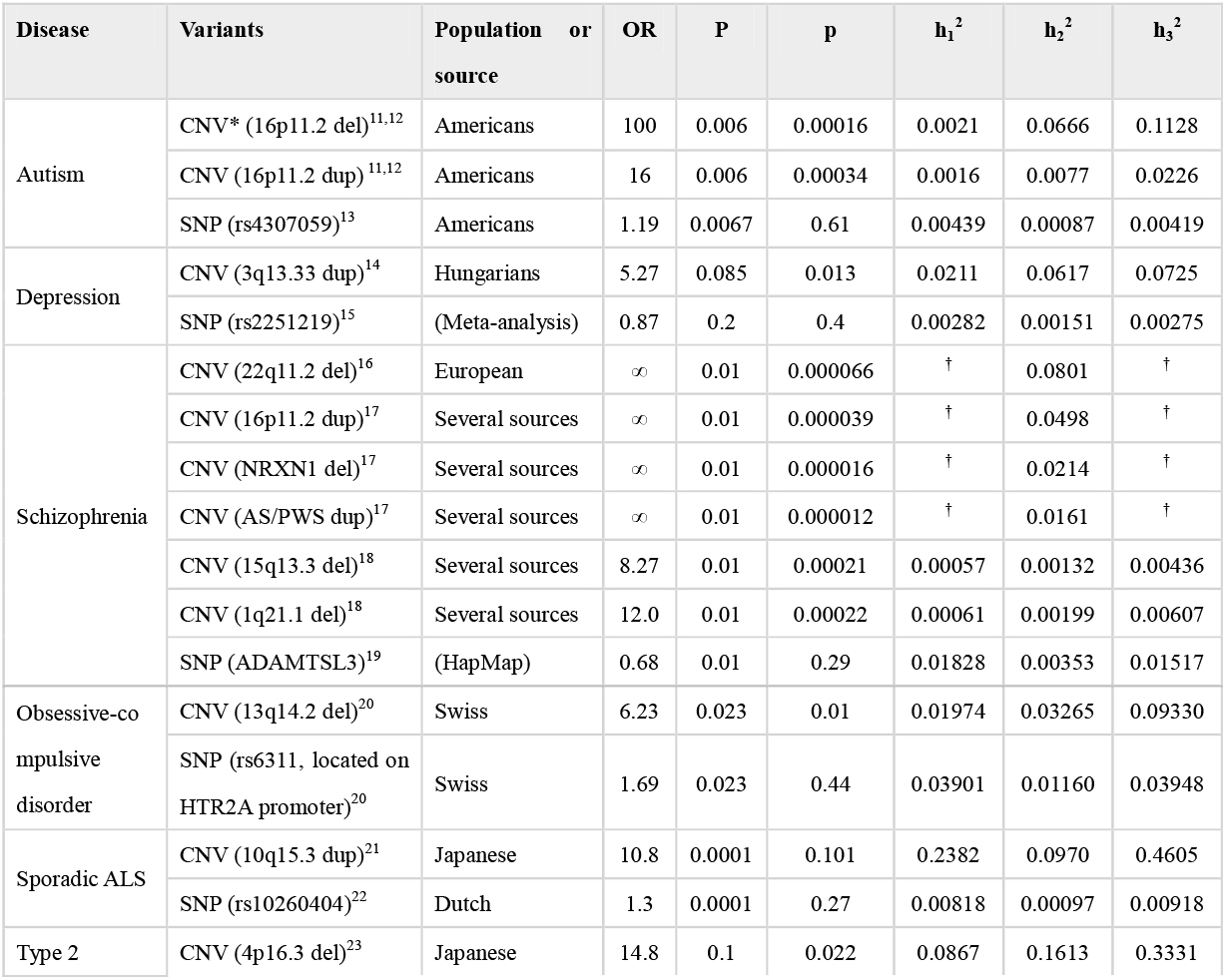
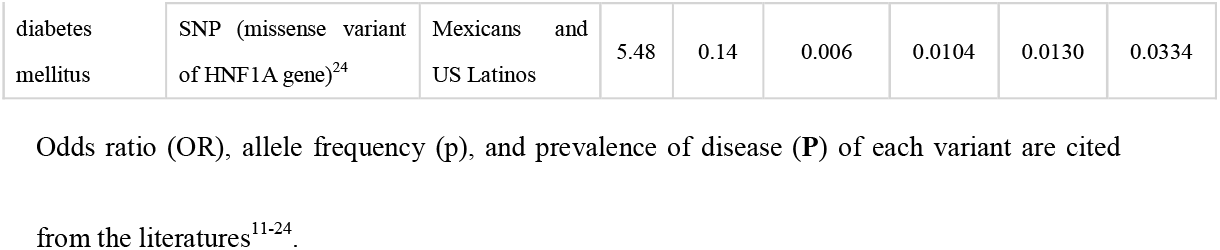
Results of a trial to calculate h^2^ by the allele distribution model (h_1_^2^), the mutation model (h_2_^2^) and the modified allele distribution model (h_3_^2^) of CNVs and SNPs using published data.

In Table 2, features of the three methods are summarized.

**Table 2.**
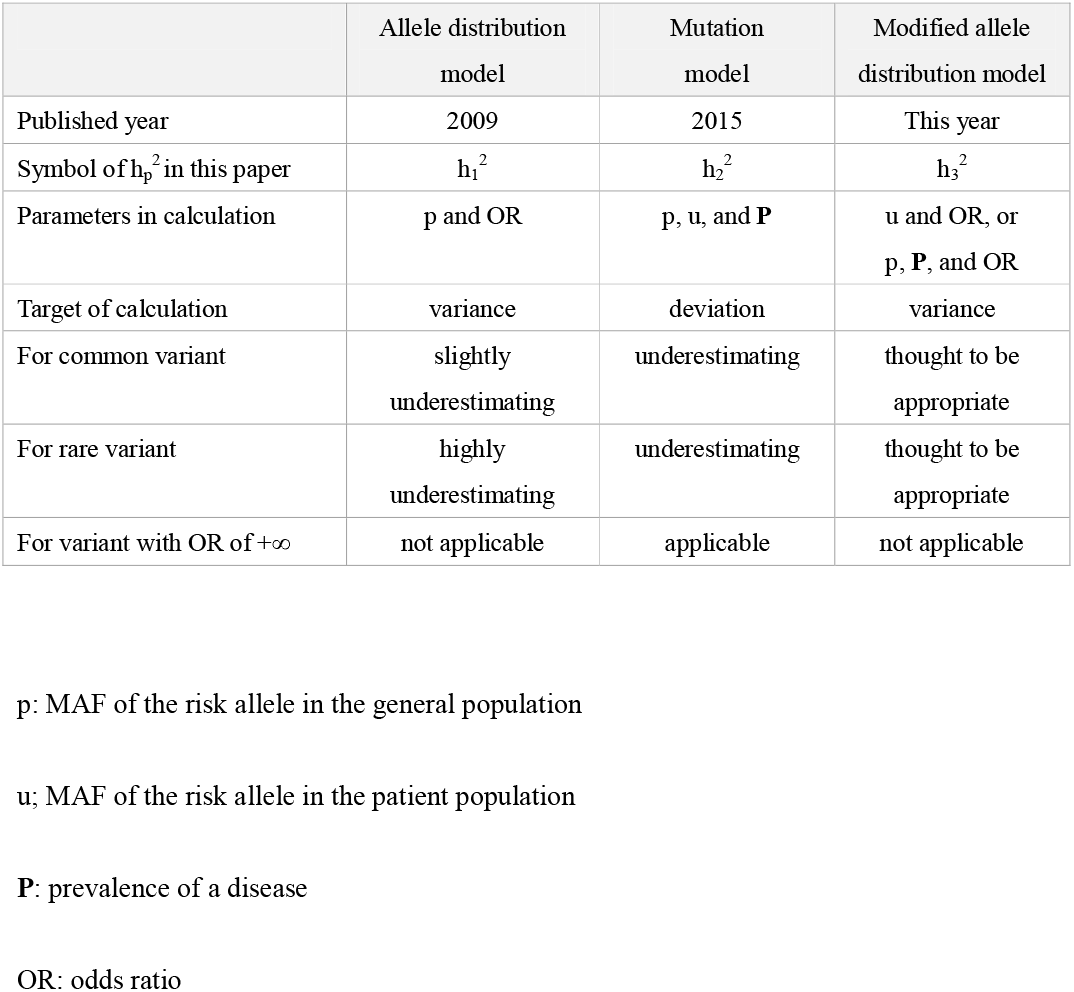
Features of three different methods to estimate h_p_^2^

|  | Allele distribution<br>model | Mutation<br>model | Modified allele<br>distribution model |
| --- | --- | --- | --- |
| Published year | 2009 | 2015 | This year |
| Symbol of $h_p^2$ in this paper | $h_1^2$ | $h_2^2$ | $h_3^2$ |
| Parameters in calculation | p and OR | p, u, and **P** | u and OR, or<br>p, **P**, and OR |
| Target of calculation | variance | deviation | variance |
| For common variant | slightly<br>underestimating | underestimating | thought to be<br>appropriate |
| For rare variant | highly<br>underestimating | underestimating | thought to be<br>appropriate |
| For variant with OR of $+\infty$ | not applicable | applicable | not applicable |

p: MAF of the risk allele in the general population
u; MAF of the risk allele in the patient population
**P**: prevalence of a disease
OR: odds ratio

## Discussion

The results show significant differences in the value of h_p_^2^ depending on the calculation method used. As shown in Fig. 1 and Table 2, the values of h_p_^2^ by three calculation methods (h_1_^2^, h_2_^2^ and h_3_^2^) show similar trends, although **P** is different. As mentioned in the derivation of the modified allele distribution model, the AF of the variant in the patient population is appropriately used to calculate the genetic variance of the variant. Accepting that formulation, the values of h_1_^2^ by the allele distribution model without the modification proposed here are inaccurately low.

Why then does h_2_^2^ take a lower value than h_3_^2^? A possible cause is as follows: h_2_^2^ is calculated with **P** and **Q** using the liability threshold model supposing that only the variant is a genetic factor for a disease. **Q** is calculated with Equation (12), which assumes that the non-risk genotype has no genetic effect. On the other hand, h_1_^2^ is calculated using Equations (17) and (18), which are derived considering all genotypes of the variant^5^. Because h_3_^2^ is calculated using Equations (17) and (18), which are applications of Equations (13) and (14), all genotypes are also considered in the calculation of h_3_^2^. Therefore, because the mutation model does not consider the contribution of the non-risk genotype of the variant to h_p_^2^, h_2_^2^ is assessed as smaller than h_3_^2^.

As shown in Figs. 1e to 1h, h_2_^2^ becomes closer to h_3_^2^ as the OR increases. In other words, at high OR, the h_p_^2^ of a variant calculated with the modified allele distribution model becomes closer to h_p_^2^ calculated with the mutation model This reflects the transition of a rare variant to a rare allele causing Mendelian disease when the effect size becomes high^11^.

h_3_^2^ calculated with the modified allele distribution model suggests that the contribution of rare variants with high ORs to h^2^ of a common disease has been generally underestimated with the original allele distribution model. Well then, is h_2_^2^ which is calculated with the mutation model meaningless? As shown in Table 2, for a variant in which the risk allele is not found in the control group, *V(g)* becomes infinite when the modified allele distribution model is used, and the non-risk allele of such a variant can be regarded as the wild type allele. I infer that reevaluation of the h^2^ is required for all the variants already identified as associated with diseases – frequent as well as rare.

To carry out such reevaluation h^2^ for a common disease using the modified allele distribution model, one must calculate *V(g)* for each variant and then estimate V_G_ as the sum of all *V(g)*s. Even when the mutation model is appropriate, the sum of (**Q** − **P**) must first be calculated for each variant, rather than using the sum of individual h_p_^2^ values (Nagao, 2015).

In conclusion, the analysis indicates that genetic variance of a variant affecting a disease should be calculated using the AF in the patient population, and as was seen for sample instances presented here, h_p_^2^ for rare variants with high ORs then is scored as significantly higher than h_p_^2^ calculated using AFs for the general population. Because rare variants show high ORs compared to common variants in average^25^, the contribution of rare variants to the heritability of common diseases is likely much larger than previously thought. Therefore, for diseases and qualitative traits, one source of “missing heritability” is likely to be the use in conventional methods of AF of a variant in the general population rather than in the patient population^5,6^. Reevaluation of h^2^ for previously identified variants and discovery of novel rare variants would thereby significantly reduce the level of missing heritability for common diseases and possibly for qualitative traits as well.

## Methods

### Derivation of the frequency of the risk genotype in the first-degree relatives of a patient

Let the frequency of the risk genotype(s) of a variant and its penetrance be X_1_ and α in the general population, respectively. X_1_ represents (p^2^ + 2pq) for a dominant risk allele; p for a co-dominant risk allele; and p^2^ for a recessive risk allele.

Suppose that this variant is the only genetic factor for a disease. Assuming that the non-risk genotype is not involved in onset, the cumulative population incidence of a disease, **I**, is represented by the following equation, including the incidence attributable to environmental factors as E:

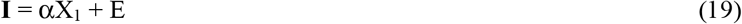

### Derivation of population attributable risk

Population attributable risk (PAR) is the proportion of the incidence of a disease in the population that is due to exposure. Table 3 shows the four basic quantities, A, B, C, and D applicable to a case-control study. PAR is represented by the following formula:

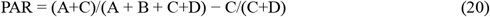

**Table 3.**
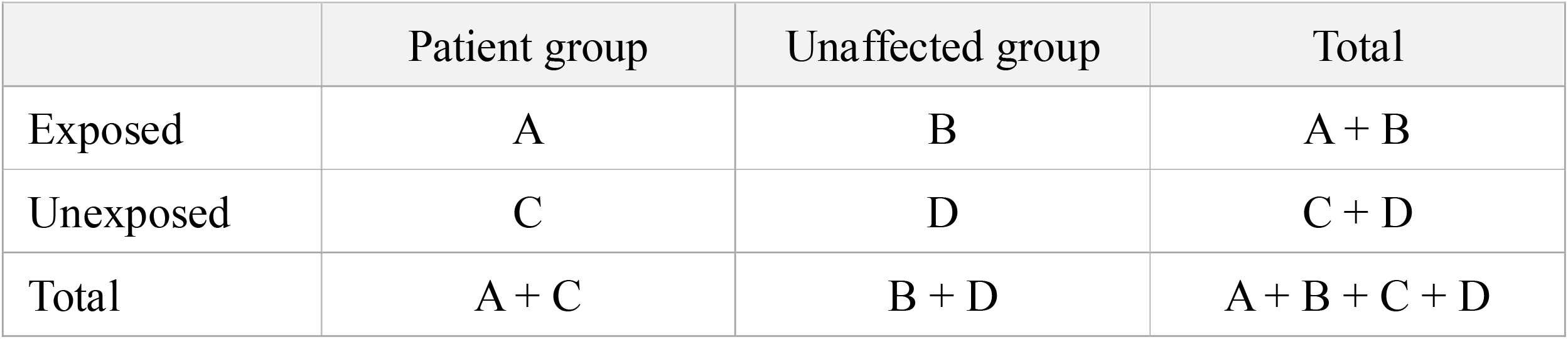
Four basic quantities in the case-control study

In Equations (6) and (7), the ratios of **P**u, (1 − **P**)x, **P**v, and (1 − **P**)y correspond to the ratios of A, B, C, and D in Table 3, respectively. Therefore, PAR is represented as following:

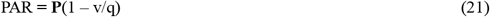

The following equation then holds:

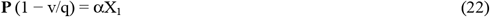

### Derivation of the frequency of the risk genotype in the first-degree relatives of a patient

Regarding the incidence in the first-degree relatives of a patient, **Q**, if the frequency of the risk genotype(s) is X_2_ and the penetrance is α’, **Q** is represented as following:

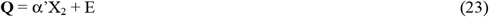

Because the relatives of the patient belong to the same population, α’ is equal to α. Then **Q** is represented as

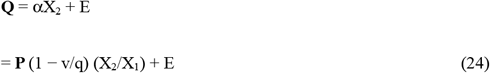

Using Equations [19], [21] and [24], **Q** is equivalent to

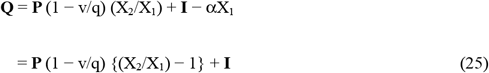

The cumulative population incidence **I** of each disease is approximated by the prevalence **P** for a chronic disease. Therefore, **Q** is represented by the following equation:

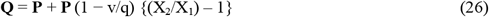

When this equation is applied to a variant under the recessive model, the heterozygote is a non-risk genotype, and the population attributable risk should be qualified^8^.

### The frequency of the risk genotype for the variant under the co-dominant model

To assess the contribution of a variant to the incidence of a disease in a population, suppose a variant, Var1, is associated with the disease. Definitions and premises are then as follows:

- p_1_ is the minor AF of risk alleles of Var1.
- q_1_ is the major AF of non-risk alleles of Var1.
- M and N are the risk and non-risk alleles, respectively.
- a, b, and c are the respective penetrance of genotypes MM, MN, and NN.

**I**, is represented by

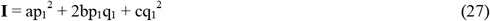

Assuming that the genetic mode of Var1 is codominant and that heterozygotes show an additive effect, then 2b = a + c. Therefore, I can be represented as

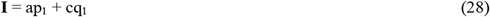

As shown in Equation (26), for the variant under a co-dominant model, the sum of the frequency of homozygosity of the risk allele and half the frequency of heterozygosity is regarded as the “frequency of risk genotype in calculation”; it is p_1_.

### Calculation of genotype frequency of risk allele for the first-degree relatives

To calculate genotype frequencies of the risk allele for first-degree relatives,

- M and N represent risk and non-risk allele, respectively.
- β is the genotype frequency of MM for the proband.
- γ is the genotype frequency of MN for the proband.
- δ is the genotype frequency of NN for the proband.

Probabilities of each genotype for offspring is as follow^8^: MM:

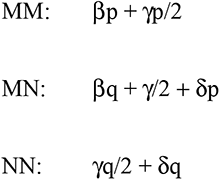

For the proband, β, γ, δ become u^2^, 2uv, and v^2^, respectively. Therefore, the genotype frequency of MM for offspring is up and that of MN is (u^2^q + uv + v^2^p). Then, for co-dominant inheritance, X_2_ is represented as follow:

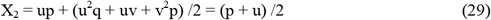

As described above, the risk genotype(s) of a variant and its penetrance, X_1_ represents p for a codominant risk allele. Therefore, Therefore, **Q** is represented by the following equation:

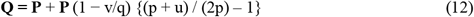

## Data Availability

All data produced in the present work are contained in the manuscript.

## Data availability

All data generated or analysed during this study are included in this published article.

## Acknowledgements

I am grateful to Dr. David Schlessinger of National Institute of Aging for valuable advice.

## Contributions

Y.N. designed the study. Y.N. is responsible for the assessment and discussion of the obtained results and wrote the manuscript.

## Additional Information

Competing financial interests: The authors declare no competing financial interests.

